# Vaccine-induced humoral and cellular immunity against SARS-CoV-2 at 6 months post BNT162b2 vaccination

**DOI:** 10.1101/2021.10.30.21265693

**Authors:** Hideaki Kato, Kei Miyakawa, Norihisa Ohtake, Yutaro Yamaoka, Satoshi Yajima, Etsuko Yamazaki, Tomoko Shimada, Atsushi Goto, Hideaki Nakajima, Akihide Ryo

**Author notes:** **Corresponding author** Akihide Ryo, M.D., Ph. D, Department of Microbiology, Yokohama City University School of Medicine.

## Abstract

To evaluate vaccine-induced humoral and cell-mediated immunity at 6 months post BNT162b2 vaccination, immunoglobulin G against SARS-CoV-2 spike protein (SP IgG), 50% neutralizing antibody (NT_50_), and spot-forming cell (SFC) counts were evaluated by interferon-γ releasing ELISpot assay of 98 healthy subjects (median age, 43 years). The geometric mean titers of SP IgG and NT_50_ decreased from 95.2 (95% confidence interval (CI) 79.8–113.4) to 5.7 (95% CI 4.9–6.7) and from 680.4 (588.0–787.2) to 130.4 (95% CI 104.2–163.1), respectively, at 3 weeks and 6 months after the vaccination. SP IgG titer was negatively correlated with age and alcohol consumption. Spot-forming cell counts at 6 months did not correlate with age, gender, and other parameters of the patients. SP IgG, NT_50,_ and SFC titers were elevated in the breakthrough infected subjects. Although the levels of vaccine-induced antibodies dramatically declined at 6 months after vaccination, a certain degree of cellular immunity was observed irrespective of the age.

## Introduction

Severe acute respiratory coronavirus-2 (SARS-CoV-2) vaccine is a powerful tool to control the coronavirus disease-2019 (COVID-19) pandemic and the mRNA vaccine BNT162b2 (Pfizer-BioNTech, USA) is reported to have an vaccine efficacy of 95% [1]. However, the long-term decline in efficacy has been a major concern regarding the efficacies of most vaccines in preventing infection; reportedly, most vaccines exhibit declining efficacies after 6 months after the vaccination [2,3]. However, the efficacy of a vaccine in preventing severe disease is reported to remain at a high level. The titer of immunoglobulins (IgG) against spike proteins (SPs) induced by the vaccine dropped to 7% in 6 months [4]. The decline in the efficacy for preventing the infection can be attributed to the decrease in antibody titer, although the prevention of severe disease cannot be explained by antibody titer alone and cellular immunity may be involved. mRNA vaccines induce not only humoral immunity but also cellular immunity [5]. We previously evaluated IgG against SARS-CoV-2 SP (SP IgG) by chemiluminescent enzyme immunoassay (CLEIA) before and up to 6 weeks after the first dosing with BNT162b2 vaccine [6]. In this study, we analyzed the SP IgG level against the original and Delta strains by CLEIA and 50% neutralizing titer (NT_50_) at 6 months after vaccination. In addition, SARS-CoV-2-specific T-cell response was evaluated by interferon γ (IFN-γ) releasing ELISpot assay to assess the status of cellular immunity at 6 months after vaccination.

## Subjects and methods

The healthcare workers at the Yokohama City University Hospital who had completed two doses of the BNT162b2 vaccine (Comirnaty 30µg, Pfizer/BioNTech, USA) given intramuscularly on the deltoid muscle (lot number: EP9605 as a first dose and ER9480 as a second dose) from March to April 2021 at 3-week interval were recruited in this study. For the 6-month post-vaccination evaluation, the blood samples were drawn at 180 ±15 days after the second dose of vaccination. The subjects’ information, including the date of birth, sex, current drinker, current smoker, comorbidities, body height, and body weight at the time of first dose of vaccination were obtained. The participants’ age and body mass index (BMI) at the time of first vaccination was then calculated.

Immunoglobulin G titer against SP and nucleocapsid (NP) IgG were determined using the commercial chemiluminescent enzyme immunoassay (AIA-CL SARS-CoV-2 SP IgG antibody detection reagent, Tosoh, Japan). Furthermore, the IgG index values between the original and Delta stain (D614G, T478K, P681R, and L452R) were measured and compared as previously described [6]. The cutoff values (1.0 index value) for NP and the SP IgG index were determined according to the manufacturer’s instructions. The neutralizing titer assay was performed using an HIV-based pseudovirus bearing the SARS-CoV-2 spike. Then, 50% neutralizing titer (NT_50_) values were calculated using the Image J software (NIH). NT_50_ values equal or less than 50 were considered negative. All samples were assayed in at least duplicate experiments. SARS-CoV-2-specific T-cell response was measured using interferon (IFN)-γ ELISpot analysis (T-SPOT Discovery SARS-CoV-2, Oxford Immunotec Ltd, UK). The corresponding blood samples were submitted for examination at SRL laboratory (H.U. Frontier Co., Ltd. Tokyo, Japan). As the specific T-cell response against SARS-CoV-2, the number of spots forming cells (SFC) per 250000 peripheral blood mononuclear cell (PBMC) reported were calculated by subtracting the number of the negative control panel from the number of SFC in Spot 1 (peptide pool for SP). SFC was multiplied by four to express the final results in 10^6^ PBMCs.

Continuous data were presented either as means with 95% CI or medians with interquartile range (IQR). Categorical data were presented as numbers and percentages. Continuous variables between the two groups were compared using the two-tailed Mann– Whitney U-test. Categorical data were compared using the Fisher’s exact test. The correlation between two continuous numbers was calculated by Spearman’s correlational analysis. A multivariable regression model was employed to investigate the association between the background variables and antibody titers. The SP IgG index values were log-transformed for analysis to remove positive skewness. Statistical analyses were performed using the Prism 9.1 (GraphPad Software, San Diego, CA, USA) and JMP Pro 15 software (SAS Institute, Cary, NC, USA). *P* < 0.05 were considered to indicate statistical significance. We previously analyzed the SP IgG index titers after 1 and 3 weeks after two doses of BNT162b2 in the other study [6]. In this study, written informed consent was obtained independently to participate in this study from each participant. This study was approved by the relevant institutional review board (approval number: B210800024).

## Results

After excluding 4 subjects who failed to submit their blood samples at the designated date and 3 subjects who were undergoing antitumor chemotherapy or immunosuppression, 98 subjects were analyzed. The subjects were of median age 43 (IQR 38–49) years, BMI of 21.8 (IQR 19.9–24.2), and included 24 men and 74 women. There were 32 (32.7%) current drinkers and 5 (5.6%) current smokers among the subjects. There were 4 patients with COVID-19 history before vaccination and 4 patients with breakthrough infection that occurred after the completion of the second dose of vaccination. The breakthrough infections were individual cases. The occupational categories were 18 physicians, 37 nurses, 20 technicians, 6 pharmacists, and 17 others.

The SP IgG index titer and GMT of all the participants are shown in Figure 1A. In the 4 breakthrough infected subjects, SP IgG at 6 months ranged from 146.1 to 459.1. In 94 subjects, excluding the 4 breakthrough infected subjects, GMT of SP IgG index titer was 47.7 (95% CI 36.8, 61.9), 95.2 (95% CI 79.8, 113.4), and 5.7 (95% CI 4.9, 6.7) at 1 week, 3 weeks, and 6 months after vaccination. The ratio of SP IgG index titer to the Delta strain to the original strain was 0.605 (IQR 0.515–0.718) after 3 weeks of 2 doses and 0.609 (IQR 0.539–0.693) after 6 months. The GMTs of NT_50_ in 98 subjects were 727.5 (95% CI 597.3, 886.1), 680.4 (588.0, 787.2), and 130.4 (104.2, 163.1) at 1 week, 3 weeks, and 6 months after the second dose of vaccination, respectively (Fig. 1B). In 94 subjects, excluding the 4 cases of breakthrough infection, the GMT of NT_50_ was 111.4 (95% CI 94.15, 131.7) 6 months after the second dose. In the 4 breakthrough infected subjects, the SP IgG index titers ranged 146.1 to 459.1, while NT_50_ ranged from 1631 to 8756. The ratio of SP IgG index against the Delta/original strains (0.902 to 0.954) was high. A correlation was noted between the SP IgG index titer and age and alcohol consumption. The GMT of SP IgG index titer among non-drinkers was significantly higher than that among current drinker {7.57 (5.89, 9.73) vs 5.34 (95% CI 3.37, 8.44)}. Multivariable regression analysis performed to exclude the confounders revealed that age and alcohol consumption were negatively correlated with the SP IgG index titer (Table 1). SP-specific T-cell count for all participants was 84 (IQR 43–188) (range: 0–700)/10^6^ PBMCs. The number of SFC in the 94 subjects without breakthrough infection was a median of 80 (IQR 40–176) (range: 0–472)/10^6^ PBMCs. A weak correlation was noted between the SP-specific T-cell response and the SP IgG index titer (r = 0.216, *P* = 0.033), albeit no correlation was noted with NT_50_ (r = 0.181, *P* = 0.076). The SP-specific T-cell response was observed in fourteen subjects who were negative for NT_50_ {48 (IQR 22– 113)/ 10^6^ PBMCs}. The SP protein-specific T-cell response did not correlate with age, gender, BMI, alcohol consumption, or smoking status (Appendix Table 1).

**Table 1.** Multivariable regression analysis of the background characteristics affecting the SP IgG index titers in subjects at 6 months after the second BNT162b2 vaccination in subjects without COVID-19 breakthrough infection.

| Background |  |  |  |
| --- | --- | --- | --- |
| factor | $\beta$ | 95% CI | *P* value |
| Age (year) | −0.0099 | (−0.018, −0.002) | 0.012 * |
| Male gender | 0.0321 | (−0.132, 0.196) | 0.699 |
| BMI | 0.0120 | (−0.008, 0.032) | 0.232 |
| Current drinker | −0.1839 | (−0.332, −0.036) | 0.015 * |
| Current smoker | −0.1179 | (−0.420, 0.184) | 0.440 |

Abbreviations: BMI, body mass index; CI, confidence interval.
\* statistically significant.

**Fig. 1.**
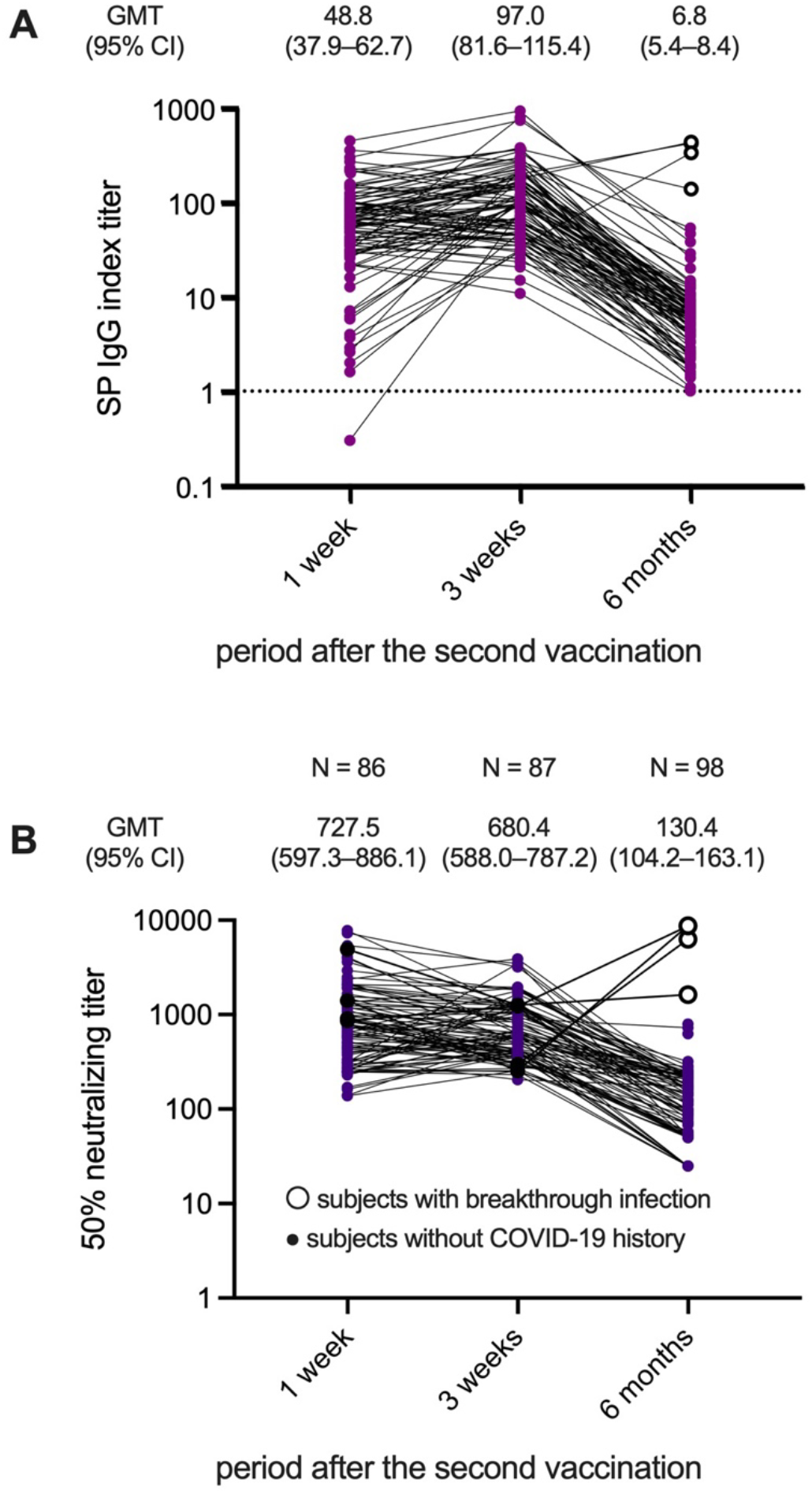
(A, B) Spike protein (SP) IgG index titer (A) and the 50% neutralizing titer (NT_50_) (B) at 1 week, 3 weeks, and 6 months after the second dose of the BNT162b2 vaccination. The white circle represents subjects with breakthrough infection after the completion of vaccination. SP IgG index 1.0 was set as the threshold of positive antibody according to the testing kit manufacturer. Abbreviations: GMT, geometric mean titer; CI, confidence interval.

**Fig. 2.**
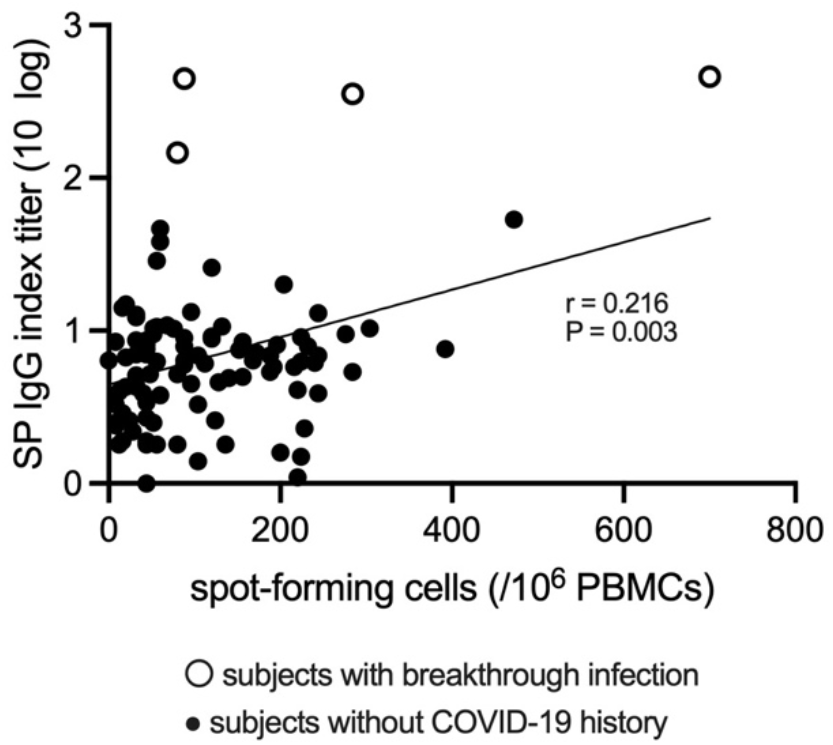
Association between several spots forming cells (SFC) using T-SPOT SARS-CoV-2 and immunoglobulin G against spike protein (SP IgG). The white circle represents subjects with breakthrough infection after the completion of vaccination. The number of SFC and SP IgG index titer showed a weak correlation. Correlation analysis was performed using Spearman’s correlation analysis, comparing 10 logged SP IgG indexes and the number of SFC. Abbreviations: SP IgG, immunoglobulin G against spike protein; PBMC, peripheral blood mononuclear cell.

## Discussions

At 6 months of receiving the second dose of BNT162b2 vaccine, SP-specific IgG decreased markedly, with a mean GMT decreasing from 95.2 at 3 weeks after vaccination to 5.7 at 6 months. A previous report showed a peak at 1 week after 2 doses and a decrease to 7% at 6 months [4]. Our data using the CLEIA method demonstrated a surprising decrease to 1/15 after 3 weeks of vaccination, and the same trend was noted for NT_50_, which also depicted a marked decrease. However, the correlation between cellular immunity assessed by SP-specific T-cell response and the SP IgG index titer and NT_50_ was weak, suggesting that cellular immunity may have a different dynamic from antibody titer. In this study, the SP-specific T-cell response in this study was 84 SFC/10^6^ PBMCs. The previously reported SP-specific T-cell response immediately after vaccination was 184 SFC/10^6^ PBMCs after two doses of BNT162b2 vaccine [7]. The ELISpot assay at 6 months in naturally infected individuals reported 97 (IQR 38–143) SFC/10^6^ PBMCs, and the SP-specific T-cell response observed in the study participants at 6 months post-vaccination was not significantly different from that observed in naturally infected individuals (P = 0.865, calculated using supplemental data from reference [8]). There seemed no difference between cellular immunity conferred by natural infection and the reduction in cellular immunity by vaccines. It has been previously reported that the antibody titer is negatively correlated with age up to 6 months after vaccination [9], but SP-specific T-cell response has not been related to age [10]. In our study, there was no correlation among age, gender, BMI, alcohol, or smoking. The decline in cellular immunity was slower than that of antibody titer, indicating it to be less negatively correlated with age, which possibly explains its long-term effect in protecting from developing the severe form of this disease.

In this study, four cases of breakthrough infection after the second vaccination were investigated. It is believed that a high antibody titer was induced in patients with breakthrough infection. Although the titer of antibodies against SP of the Delta strain was 60% of that against the SP of original strain in the patients without breakthrough infection, the ratio of anti-Delta/anti-original antibody titers was as high as 90% in the patients with breakthrough infection. In Japan, the Delta strain has been the predominant strain since April 2021 [11]. This study has several limitations. For instance, the data on SP-specific T-cell response immediately after the vaccination was lacking and the sample size of the cohort was small. In addition, generally, cellular immunity is weakened in elderly and immunosuppressed individuals. However, our study demonstrated that antibody titers do not necessarily correlate well with cellular immunity, indicating that the dynamics of cellular immunity are different from those of humoral immunity and suggesting that the evaluation of cellular immunity is warranted for long-term evaluation of the efficacy of SARS-CoV-2 vaccines.

## Data Availability

All data produced in the present study are available upon reasonable request to the authors.

## Author contributions

HK contributed to the study design, data collection, statistical analysis, and interpretation of data, as well as the drafting and editing of the manuscript. SY, EY and TS contributed to the study design and data collection. KM, NO, and YY performed the laboratory tests. TM, AG, HN, and AR contributed to the study design, data collection, supervision of the analysis, and preparation of the manuscript. All authors made critical revisions to the manuscript for important intellectual content and approved the final manuscript. All authors meet the ICMJE authorship criteria.

## Acknowledgments

The authors thank all the participants and medical staff for their participation and assistance in the study. This work was supported by the Japan Agency for Medical Research and Development (AMED) under Grant Number: JP20he0522001. The authors would like to thank Enago (www.enago.jp) for the English language review.

